# Implementation of a shared decision making process for severe stroke-a mixed methods study

**DOI:** 10.1101/2024.02.21.24303139

**Authors:** Akila Visvanathan, Sarah Morton, Allan Macraild, Polly Black, Sophie Gilbert, Mark Barber, Martin Dennis, Richard O’Brien, Gillian Mead

**Author notes:** <u>Acknowledgements</u>. We thank Jessica Crossan, Mairi MacDonald, Sarah Risbridger (research nurses) for recruiting patients. This work was supported by Edinburgh and Lothian Health Foundation Reference 1339. Ethics approval was given by Scotland A research ethics committee (21/SS/0044). Address for correspondence, Western General Hospital, Crewe Road Edinburgh EH4 2XU.

## Abstract

**Background:** Clinical decisions made early after stroke can make the difference between survival with disability or death. We aimed to develop, implement and evaluate a new Shared decision making (SDM) process for severe stroke into a regional 36 bedded stroke unit.

**Methods:** We developed the process through four coproduction workshops, attempted its implementation then its impact on death at 6 months, discharge destination and tube feeding. We also explored patients, families and staff views about SDM.

**Results:** Eleven people (staff and people with lived experience of stroke) attended the first co-production workshop, eight the second, seven the third and six the fourth. The new SDM process incorporated Tailored Talks (a digital platform with information about stroke and its prognosis) and an implementation plan (including staff training). We implemented this process on 1^st^ August 2022.

Only 8/1020 patients received Tailored Talks (4 before and 4 after implementation). For the entire group there was no change tube feeding, discharge destination or death. The proportion of people with severe strokes dead at six months was higher before implementation. Twenty-one patients or family members provided views about SDM quality, but the sample size was too small to draw conclusions. Staff interviews suggested that insufficient time, lack of a ‘human touch’ and inadequate leadership explained the lack of implementation.

**Conclusion:** Our co-produced SDM was not effectively implemented into a stroke unit and there was no change in the use of tube feeding or death in 1020 patients.

**Key points:** Shared decision making after severe stroke is complex.

A co-produced new process for shared decision making after severe stroke was not effectively implemented into clinical practice.

There was no change in tube feeding, death or institutionalisation.

## Background

Stroke is the second leading cause of death [1]. Around 50% of patients are left disabled [2]. Stroke can cause from mild, quickly resolving, neurological deficits to severe persistent life-threatening deficits. After severe stroke, treatment decisions may determine whether the patient survives with severe disability or dies [3].

Because stroke is a sudden event, patients, families and caregivers are often unprepared for making treatment decisions. Patients may not have capacity because of aphasia, cognitive impairment or impaired consciousness [4–6]. Patients and their families may be in a state of shock; and the likely extent of recovery is often uncertain [7].

Shared decision making (SDM) is important because patient involvement is a fundamental right. Patients generally want information about their health condition and want to take an active role in decision making [8]. After severe stroke, patients and relatives need emotional support and prognostic information, even though outcome is difficult to predict. Health care professionals need to deal with uncertainty, and balance hope with realism [9]. Patients report that they do not always receive the opportunity for SDM after stroke [10]. The quality of SDM and quality of life after stroke are top priorities for future research in severe stroke [11].

A 2018 Cochrane review of SDM in a range of patient groups found 87 studies, but the evidence was of low certainty [8]. To the best of our knowledge there is just one trial of SDM in severe stroke, in which the feasibility of a Neurological Intensive Care Unit paper-based decision aid, for people with severe acute brain injury and stroke in the US was tested in 41 patients and 66 surrogate decision makers. [12,13]. The decision aid was feasible and well received.

To improve SDM after severe stroke in the UK, ‘Tailored Talks’ was developed. This is a digital communication platform using Powerpoint slides that facilitates tailoring, structuring and sharing of only relevant information about stroke with patients and families [14].

Our primary aim was to co-develop and embed a process for SDM for severe stroke into the stroke service at one acute hospital site, using Tailored Talks as the information source about stroke.

Our secondary aims were to evaluate whether the new process was effectively implemented, to explore whether it was associated with changes in processes and outcomes (death, discharge destination, use of feeding tubes), to evaluate the views of patients, family and staff about the quality of SDM both before and after implementation of the new process, and explore whether patients/families’ preferred outcome (death/severe disability) at baseline matches the actual outcome at 6 months.

## Method

Ethical approval was granted (see title page for committee). We used mixed methods: a) coproduction (months 1 to 4), b) implementation (month 6 onwards), c) audit (months 1-12), d) questionnaires (months 3-9) and e) qualitative interviews (months 6-12) with patients and relatives, and a focus group with staff.

### Co-production

Co-production is a collaborative research approach that involves multiple stakeholders underpinned by three principles: (i) a structured, participatory approach designed to actively engage participants to contribute; (ii) ensuring all participant voices are heard, opinions evaluated, and appropriately acted on, and; (iii) encouraging all participants to actively contribute to the development of the SDM process for embedding Tailored Talks into practice. Our coproduction group included 13 participants (stroke survivors, relatives, and stroke care professionals from a range of disciplines/seniorities) recruited through stroke charities and through professional networks.

Participants were invited to one of two introductory workshops. These were followed by four co-production workshops, each lasting about an hour, facilitated by at least two researchers (SM, AV and/or, AM), hosted online (due to Covid 19 restrictions) using NHS Scotland National Video Conferencing service between 18 January 2022 and 24^th^ May 2022. Of the 13 participants recruited, eleven of these attended the first workshop, eight the second, seven the third and six the fourth workshop.

The topics covered in each workshop were a) overview of the aims b) how to provide tailored information about prognosis c) how to elicit family/patients views and d) how to implement the new process. Participants were invited to consider different intervention functions (education, persuasion, incentivisation, coercion, training, restriction, environmental restructuring, modelling and enablement) in their appraisal of Tailored Talks and its role in a SDM process. During the workshops, the APEASE (Acceptability, Practicability, Effectiveness, Affordability, Side-effects and safety) criteria were considered [15]. Workshops were recorded using an encrypted audio recorder and transcribed verbatim into Word documents. After each co-production workshop, transcripts were imported into NVivo v11 for thematic analysis and coding by two researchers [AV, SM]. In addition to mapping the data to APEASE criteria, we also used a thematic approach because of the rich holistic data obtained. Results from the first workshop informed the development of materials and discussions at the second workshop, and so on.

### Audit

Between 1^st^ February 2022 and 31^st^ January 2023 (months 0 to 12), we extracted data on death, and place of discharge for all patients with acute stroke seen in our hospital from the Scottish Stroke Care Audit. In order to identify severe stroke (National Institute of Health Stroke Score (NIHSS) of 15 or over), clinical staff seeing patients with acute stroke agreed to record NIHSS for all patients seen during the study period. For this specific project, the audit coordinator also extracted data from the medical records on the total NIHSS score recorded by the admitting clinicians, the use of feeding (nasogastric and percutaneous gastrostomy) tubes, and the documentation of Tailored Talks (as an indicator of implementation of the SDM process).

### Questionnaires

Three months before implementation of the new process (1^st^ May 2022) and for three months afterwards, potential participants (acute severe stroke with NIHSS ≥ 15) were identified by the research team in collaboration with ward staff. If an NIHSS had not been performed by the clinical staff, the research team approached patients who appeared clinically to have had a severe stroke; and then the project PI (GM) calculated retrospectively NIHSS from information in the medical records for those patients recruited. Patients with capacity and next-of-kin were approached directly. If patients did not have capacity, only the next of kin was approached. We expected to recruit 100 participants over six-months, assuming that a quarter of the ∼1000 patients admitted per year would have had a severe stroke.

The four-item SURE test (4 items, each with yes/no responses) [16], which is a short version of the decisional conflict scale, and the three-item CollaboRATE measure to assess the perception of being informed and involved in decision-making steps [17], were completed face-to-face or by telephone by research nurses at baseline, weeks 2, 4 and 8. These time points were chosen because key decisions and advance care plans are often made around these times (e.g. hyperacute care, fluids, feeding tubes, ’Do not attempt cardiopulmonary resuscitation’, and ‘escalation’ to Critical Care, antibiotics or not for infection).

At baseline research nurses also assessed the simplified modified Rankin score (smRS) [18], and asked two open ended questions a) ‘If your (or your loved one’s) stroke was so severe that you (they) could no longer look after themselves and require care in a nursing home, what would be preferable to you (or your loved one): Dying comfortably from the stroke in hospital’? ‘Dying at home after a discharge for palliative care’ or ‘Surviving with disability but needing long-term care in a nursing home’? and b) ‘As you (or loved one) are now, would you prefer ‘Dying comfortably from the stroke in hospital’? ‘Dying at home after a discharge for palliative care’ or ‘Surviving with disability but needing long-term care in a nursing home’.

At 6 months, we obtained data on the actual outcome (death/institutional care) and completed on the telephone the smRS and asked about specific abilities (Walk (yes/No); Talk (yes/no); Eat normally (yes/No) and the anxiety/depression from the Euroquol 5D 5 level.

### Qualitative interviews and focus group

To obtain in-depth data about the quality of SDM, AV conducted five telephone interviews between 16/11/2022 and 25/11/2022 with one patient and four bereaved relatives. Guided by the Consolidated criteria for Reporting Qualitative research [19], two reviewers (AV and SM) independently coded transcripts using NVivo and performed thematic analysis. Ideally, we would have recruited more patients but we were constrained by resources.

AV and AM conducted a staff focus group on 6/12/2022, recruited in response to invitation posters in staffrooms. One research nurse and one physician associate attended. The discussion was audio-recorded, transcribed, coded using NVivo and one researcher (AV) performed thematic analysis.

## Results

### Co-production

The feedback obtained from workshops 1 to 4, ideas for implementation and what aspects of the implementation plan could be put into practice is shown in table 1.

**Table 1.**
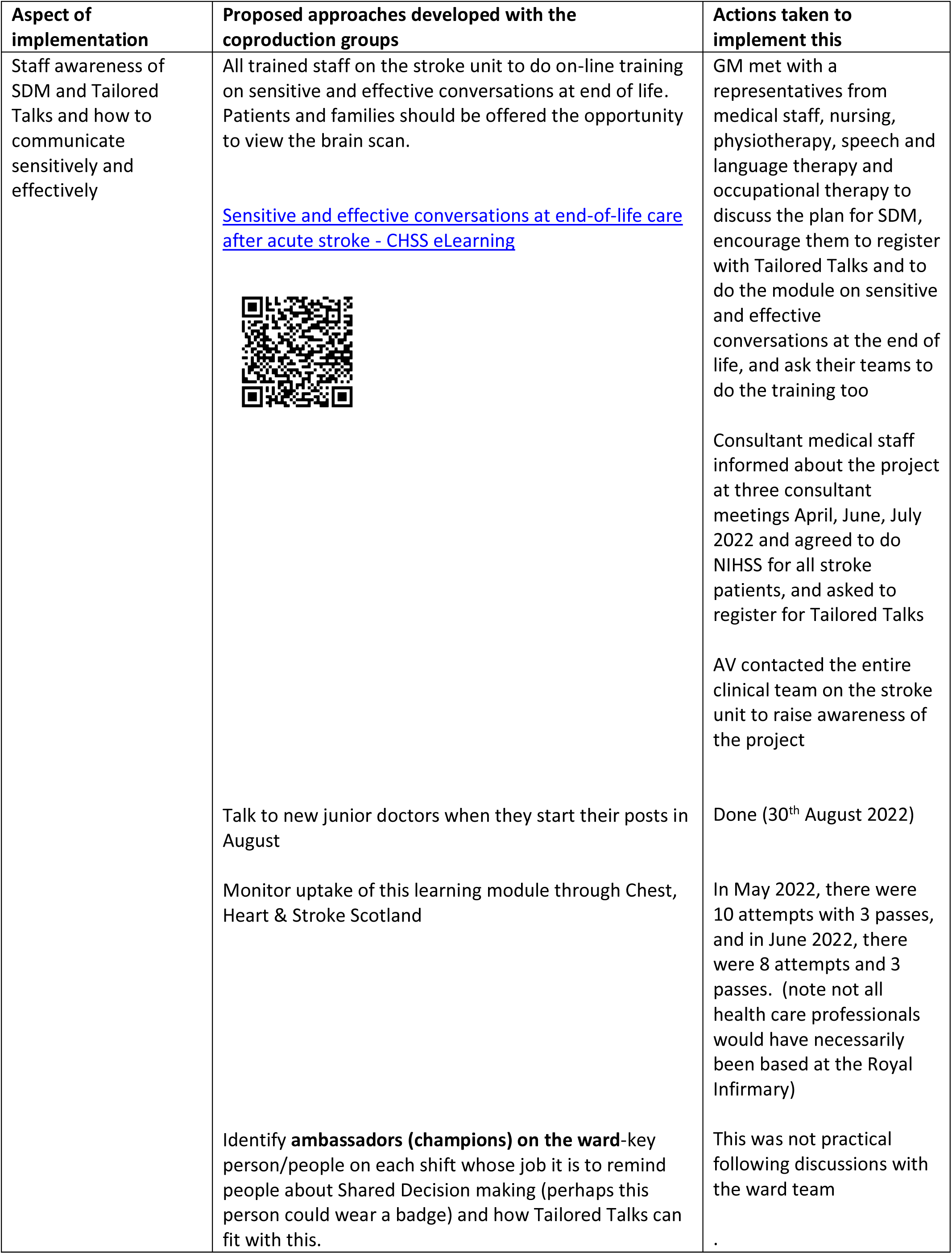
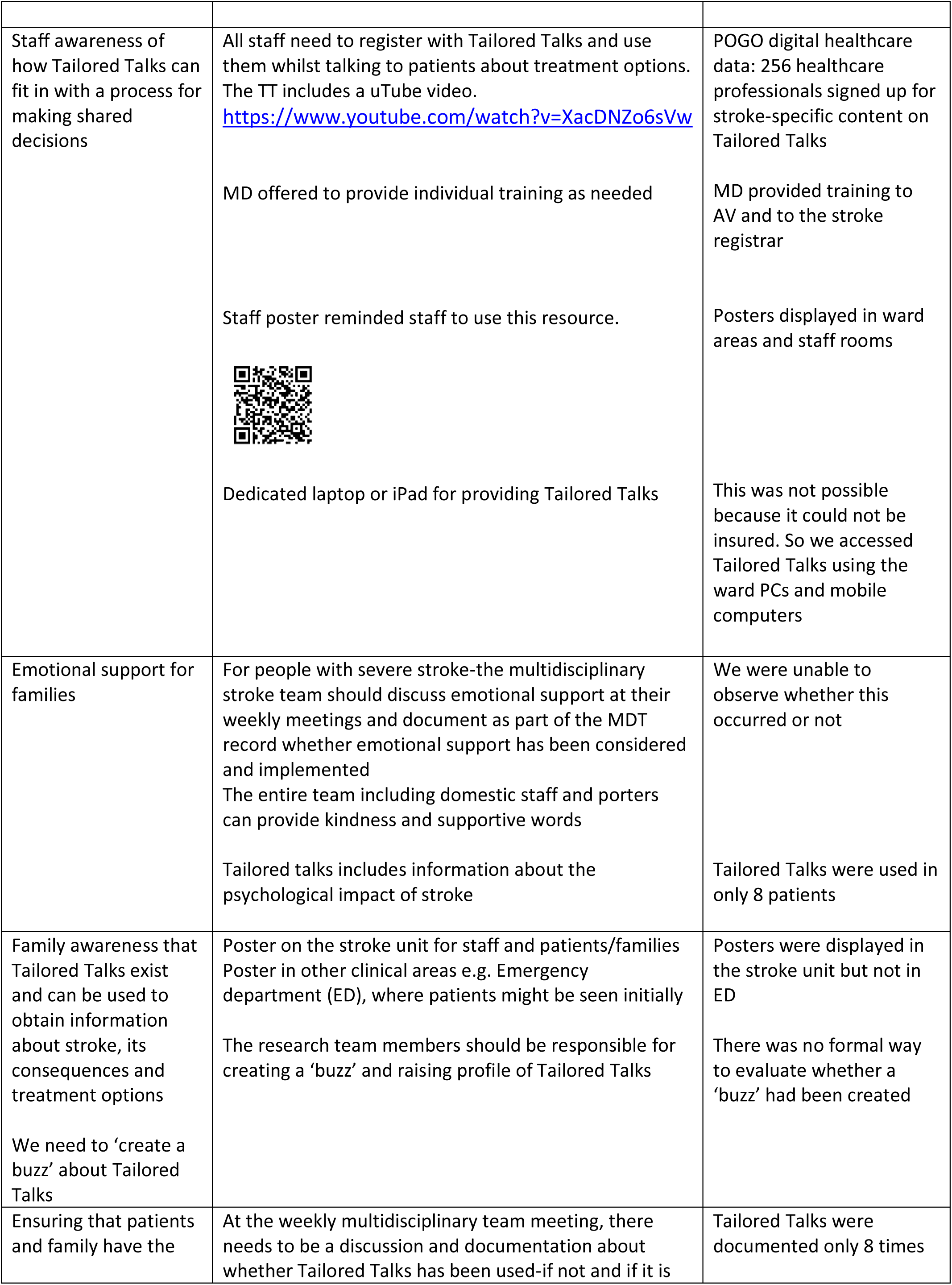
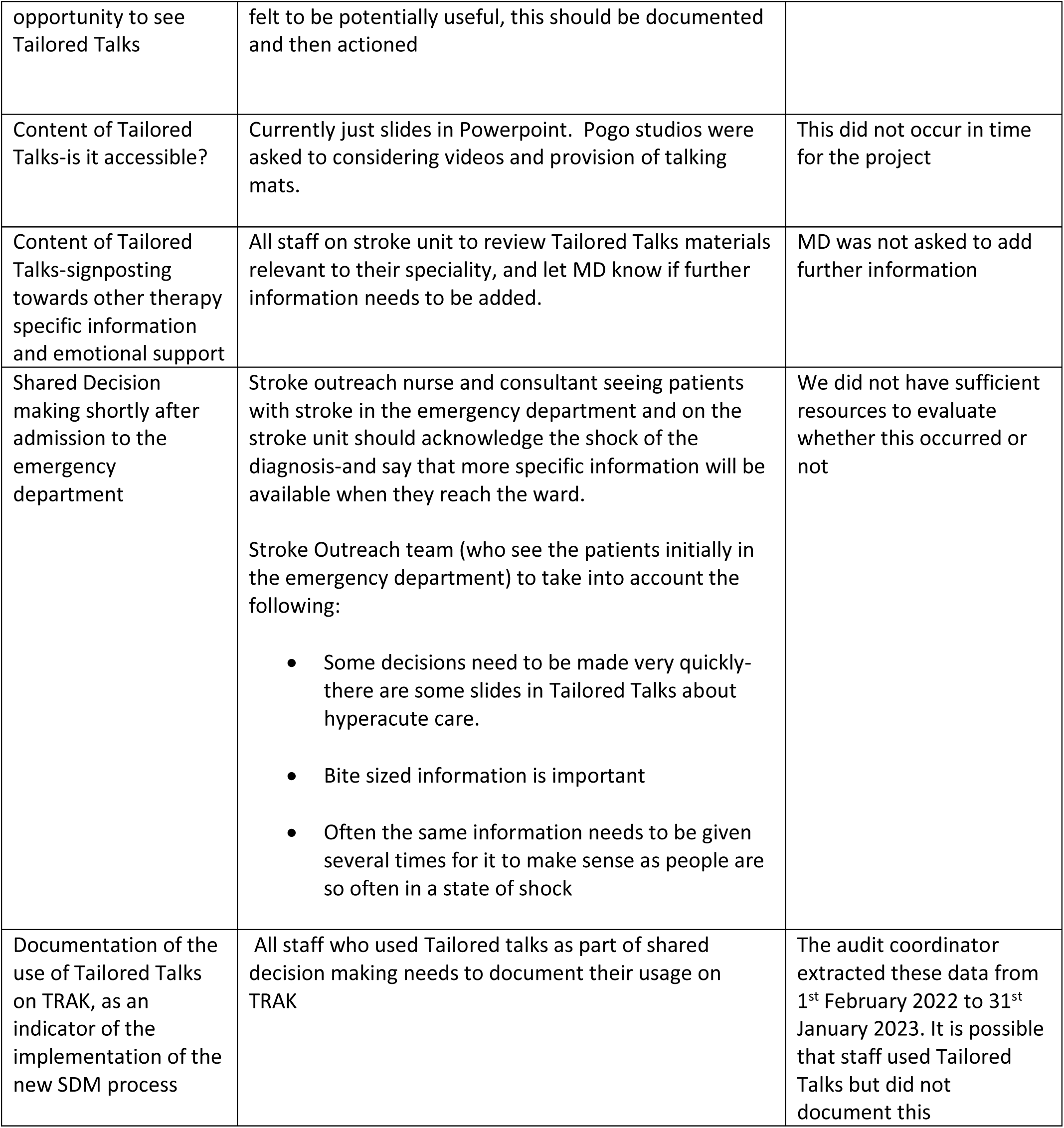
Outcome of the coproduction group and implementation.

The implementation plan was registered with our department’s Quality Improvement lead prior the official implementation date of 1^st^ August 2022. Preparatory training in Tailored Talks had been provided to staff before implementation.

Table 1 about here

Mapping of the feedback according to APEASE criteria are shown in Table 1 (appendix).

### Audit

From 1^st^ February 2022 to 31^st^ January 2023, 1020 patients (502 pre- and 518 post-implementation of the SDM process) with a diagnosis of acute stroke were admitted, mean age was 73 (SD 15) and 496 (48.6%) were female. We used an iterative quality improvement methodology to increase the proportion of patients with a documented NIHSS, but improvements were not sustained (figure 1). The overall proportion having an NIHSS assessed at admission by the clinical team was 581 (57%); of these 143 (24.6%) had a NIHSS of ≥ 15. For the entire group, there was no difference tube feeding during admission, death or institutionalisation at 6 months, before and after implementation (table 2) (Chi-squared tests). The low rate of admission to institutional care from our ward was because disabled patients were often referred on for rehabilitation in other hospitals. We did note a statistically significant reduction in death at 6 months for severe strokes-but the clinical significance of this is uncertain.

**Figure 1.**
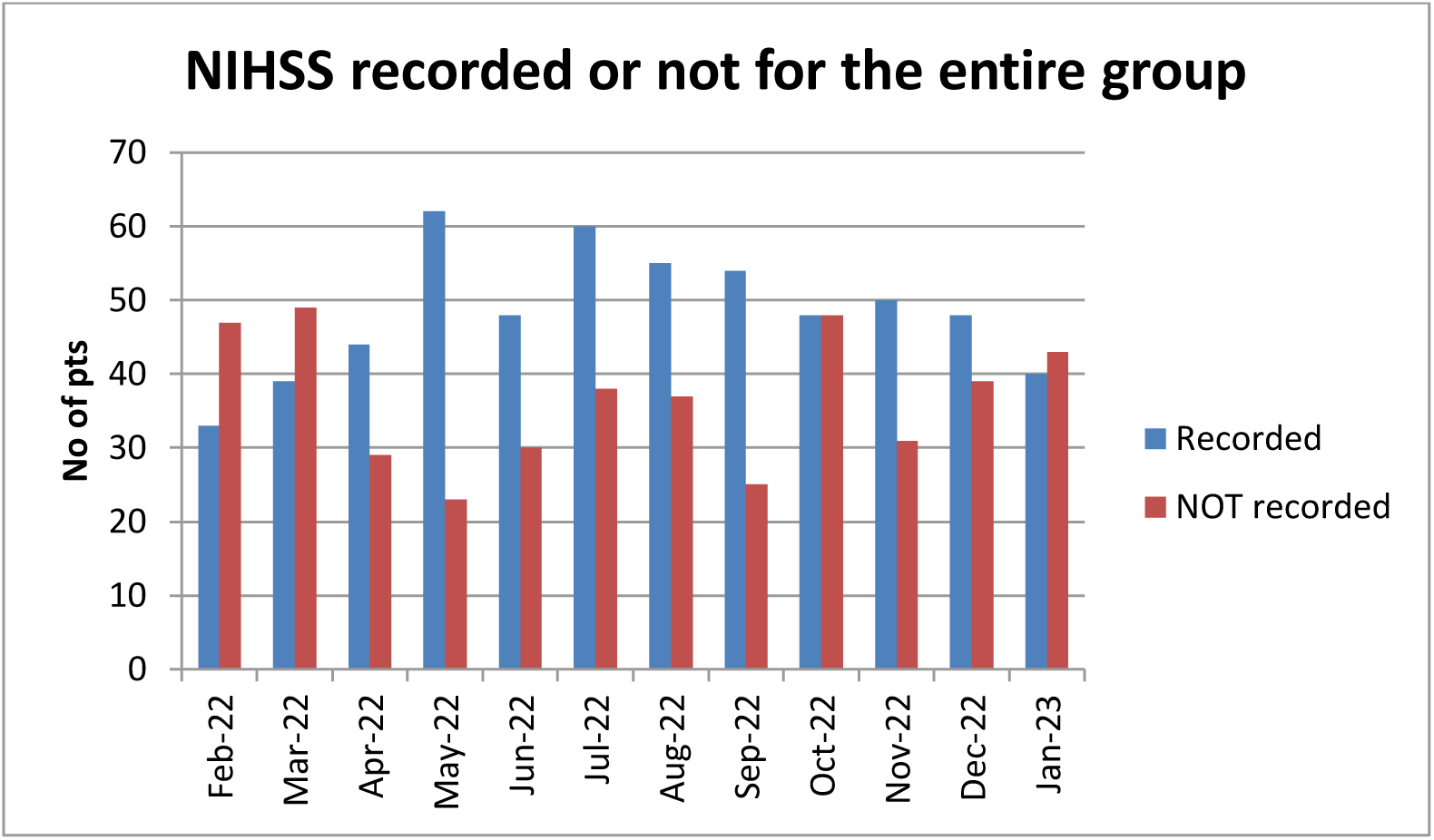
Rate of completion of the National Institute of Health Stroke Scale

**Table 2.** Process of care and outcome before and after implementation of the new shared decision process.

| Parameter | Pre implementation |  | Post implementation |  |
| --- | --- | --- | --- | --- |
| | All strokes<br>(n=502)<br>N (%) | NIHSS $\geq 15$<br>(n=68)<br>N(%) | All strokes<br>(n=518)<br>N (%) | NIHSS $\geq 15$<br>(n=75)<br>N(%) |
| Number dead at 6 months after stroke onset | 345 (69%) | 45* (66) | 352 (68) | 37 *(49) |
| Discharge destination from stroke unit | 1 (0.2) | 0 (0.0) | 3 (0.6) | 2 (2.7) |
| Feeding tube-NGT | 86 (17.1) | 21 (30.9) | 87 (16.8) | 25 (33.3) |
| Feeding tube-PEG | 3 (0.6) | 2 (2.9) | 5 (1.0) | 1 (1.3) |
| Feeding tube-both NGT and PEG | 3 (0.6) | 2 (2.9) | 5 (1.0) | 1 (1.3) |
| Tailored talks documented during hospital admission | 4 (0.8) | 2(2.9) | 4 (0.8) | 2 (2.7) |

Note-status (dead or live) for 13 patients in the first six month period and for 10 patients in the second six month period is unknown for various reasons including having moved out of the area
\*The only statistically significant difference between the first and second six month period was for higher % of severe stroke dead at 6 months in the first six month period (Chi square 4.12, p=0.04)

Figure: Documentation of NIHSS over the study period-about here

### Questionnaire data

Between 1^st^ May and 31^st^ October 2022 the research nurses identified 78 potentially eligible patients by discussion with the clinical staff; of these 37 had an NIHSS <15; five died before they could be recruited, five had no capacity, no next of kin or were not proficient in English, four declined, two were moved to another hospital/nursing home before they could be recruited and four could not be reviewed after initial contact.

Of the 21 patients (14 women, 7men), mean age 80 years old (range 41 -95) who were recruited, one was lost to follow-up before any assessments could be done. Of the remaining 20 patients, surrogate responses were obtained from next of kin/family for 18 and only two patients could answer the questions for themselves. Median NIHSS (measured at the time of stroke or calculated from record review [20]) was 23 (range 15-34). The preferred outcome at baseline is shown in table 3. At 6 months, 14 had died, three were in a nursing home and the three had been lost to follow-up.

**Table 3.** Preferred outcome at 6 months (hypothetical situation and as the patient was at the time of the interview, provided at baseline.

|  | Data (n=21) |  |  |
| --- | --- | --- | --- |
| Question | Death in hospital (n) | Survival with severe disability (n) | Lost to follow-up (n) |
| 'If your (or your relative's) stroke was so severe that you/they could no longer look after yourself/themselves and need care in a nursing home, what would you prefer?' | 17 | 3 | 1 |
| 'As you are (or as your relative is) NOW, what would you prefer?' | 13 | 7 | 1 |

Of the 14 participants who had died, only one had stated at baseline that they would rather survive as they were at the time than die in a nursing home; and all 14 said that they would rather die if they developed severe disability.

Table 3 summarises the responses obtained at time of consent with respect to the two questions asked.

Table 4 shows the CollaboRATE and SURE responses. We cannot draw any conclusions about participant perceptions before and after implementation because of the small sample size, or about functional status at 6 months.

**Table 4.** SURE and CollaboRATE-5 scores (Mean and standard deviation*) SD calculated using www.calculator.net/standard-deviation-calculator.html and set at ‘sample’

|  | Mean (SD) and number of respondents(N) |  |  |  |
| --- | --- | --- | --- | --- |
|  | Baseline | Week 2 | Week 4 | Week 8 |
| CollaboRATE<br>(maximum possible score is 12) | 9.6 (12.3)<br>N=20 | 6.8 (20.2)<br>N=11 | 7.4 (15.0)<br>N=7 | 5.8 (12.2)<br>N=5 |
| SURE (maximum possible score is 4) | 3.7 (1.7)<br>N=20 | 2.5 (3.3)<br>N=11 | 2 (3.7)<br>N=7 | 3 (3)<br>N=5 |

### Qualitative interviews

Five participants were interviewed (one patient, age 73 and four relatives of deceased patients (ages of deceased patients were 89, 83, 63 and 91). Full quotes are available on request.

Three main themes were drawn from these interviews.

1. Experience of stroke and stroke care

Some participants were complimentary of the care that they received on the ward. Value was placed on seeing brain scans and having things explained. Some participants could not remember what had happened in hospital. The shock of having a stroke, and the fear of recurrent stroke, was reported.

2. Diagnosis and discussions about stroke and treatment, involvement in decision making

There was variability in how different participants felt they had been involved in discussions about the diagnosis, prognosis and management options. Some reported that they had either not been in a position to make choices and/or ask questions about treatment (due to shock of the diagnosis or being too ill) or had been unaware that there were choices to be made.

3. Provision of information

Four participants reported how they valued information from health care professionals, and that viewing the brain scan was helpful to their understanding, especially when the prognosis of the patient was poor. Participants reported varying needs for the amount and type of information e.g. paper leaflets or various online sources. Some participants felt that information should be given after the initial shock of stroke had lessened.

### Tailored talks as a mode of information provision

One participant who had received Tailored Talks said she had been shocked to hear about the prognosis so early after stroke. The participants who did not receive Tailored Talks felt that it might have been useful.

#### Focus group

Five participants were initially recruited but only two could attend due to clinical service pressures. AV analysed the data. Full quotes are in table 3 (appendix). There were two main themes:

1. Experience of Tailored talks and its use Participants felt that it was a good source of information and as an educational resource, and had previously used it for ‘’low stake conversations’’ e.g. medication discussions but not ‘high stake’ discussions e.g. tube-feeding or end of life care.
2. Barriers to TT use. The reported barriers were insufficient time on the busy ward, the material was not ‘patient friendly and was ‘too medical’, using the material meant that eye contact and a ‘human touch’ were lost, and there had been no consultant leadership.

## Discussion

Despite using coproduction methodology to develop a new SDM process and implementation plan, our audit of >1000 patients suggests that the process was not effectively implemented and there were no changes in tube feeding, or death/institutional care at 6 months for the entire group. Our SDM process incorporated all the key elements recommended by the American Heart Association for cardiovascular SDM except for ‘decision coaches’ [20]. However, when implementing the SDM process we were not able to identify ‘champions’ on the ward, or provide iPads, as advised by our coproduction group. The staff focus group indicated that there had been insufficient time to use Tailored Talks, that the materials were not ‘patient friendly’ and that there had been a lack of consultant leadership. One relative who did see the Tailored Talks reporting feeling shocked at hearing the prognosis so early after stroke. Although we noted an apparent fall in 6 month case fatality for those with severe stroke after the implementation date, it’s unclear whether this was related to the SDM process.

Our study was designed pre-Covid-and the start date had to be delayed to February 2022. During our study, treatment escalation plans were being implemented in our health board for all patients and embedded into the electronic medical records. Thus staff may have felt that they were already practising SDM and did not need a new process.

We recruited only 21 patients to the questionnaire study; this was probably because only just over half of patients had an NIHSS recorded by the clinical staff, some patients died before they could be consented and some declined. We demonstrated feasibility of the SURE and CollaboRATE questionnaires; the baseline scores were high, but because the sample size fell over time we cannot comment on longitudinal changes. There were insufficient data to determine whether the scores were different pre-and post-implementation. Of the 14 patients who had died by 6 months, death had been their preferred outcome at baseline.

Our qualitative interviews with family members revealed themes consistent with previous studies (experience of stroke and stroke care, diagnosis and discussions about stroke and treatment, involvement in decision making and provision of information [4,6,7]. The range of participants’ responses were wide, perhaps reflecting a wide range of values, beliefs, and prior experiences (of medical care and illness), differences in stroke characteristics (e.g. severity and neurological deficits), and variation in how different health care professionals approached SDM.

Had the new process been successfully implemented into practice, our next step would have been a feasibility randomised trial, likely a step wedged design. Instead, consideration needs to be given to new literature published since we first designed this study, to decide whether we should refine our SDM process and attempt implantation again, either in the same ward or a different stroke service. Tailored Talks are being used in our health board to counsel patients about anticoagulation for atrial fibrillation, for post-Covid 19 care and to recruit patients to a platform trial of intracerebral haemorrhage [21]. Experience in these areas will inform our next steps.

In summary, our new co-produced process for SDM after severe stroke incorporating Tailored Talks was not effectively implemented into practice and there was no change in tube feeding or death. The CollaboRATE and SURE questionnaires were feasible for assessing SDM after stroke.

## Data Availability

All data produced in the present work are contained in the manuscript

## Acknowledgements-

We are grateful to patients and families who participated, the members of the Coproduction group, the clinical staff involved in the management of the patients with severe stroke, and the research nurses (who are listed on the title page).

