## Supplemental tables for "Implementation of a shared decision making process for severe stroke-a mixed methods study"

**Appendices**

Table 1 Workshop findings mapped to APEASE criteria

| **Workshop number** | What currently happens in SDM in severe stroke?<br>What is good about the current process? What could be improved?<br>What does current research tell us? |
|---|---|
| 1 Overview of aims of the co-production | INFORM |
| | • Introductions<br>• Discuss co-production workshop(s) aims and timeline, and agreement on how the group(s) can work effectively.<br>• Reminder of the aims of the project as a whole and the specific focus of including what SDM is.<br>• Presentation of evidence from our previous research. Sharing existing on-line materials that are used to support SDM<br>• Presentation of evidence from our audit of documentation of the process of SDM. |
| | KNOWLEDGE |
| | • Carry out 'character profile' and 'character journey' activities to gather knowledge about who the users of the intervention will be and what is important to them.<br>• Carry out 'asset mapping' activity to gather knowledge about what the group members already do to facilitate SDM after severe stroke<br>• Work out what training for staff might be needed |
| | EVALUATE |
| | • Summary of workshop led by facilitator with group members invited to contribute (including feedback and questions), outline next steps and date of next meeting. |
| 2 Designing a SDM tool-information for families and patients which includes information about stroke contained in | INFORM |
| | • Reflections and discussion of key points identified following workshop 1.<br>• Presentation of relevant Tailored Talks materials.<br>• Prediction of recovery of 'specific abilities'<br>• Identify how to improve/change these materials |
| | KNOWLEDGE |

| | |
|---|---|
| ‘Tailored Talks’ | • Using persons derived from workshop 1 (‘character profile’ activity (1) and ‘problems and solutions’ identified by the research team from ‘character journey’(2) and ‘asset mapping’ (3) activities ask participants to complete ‘priority matrix’ (4) worksheet<br>• Complete ‘opportunity card’ (5) activity to allow group members to suggest their idea(s) for improving the Tailored Talks. |
| | EVALUATE |
| | • Summary of workshop lead by facilitator with group members invited to contribute (incl. feedback and questions), outline next steps and date of next meeting. |
| 3. How can we elicit patient and family views, beliefs and values? Would a checklist of topics to be covered, be useful? How can we facilitate nurses, junior doctors and senior doctors to elicit such conversations? What training is needed? | INFORM |
| | • Presentation of evidence from our previous research including an audit of communication around the time of death on a stroke unit.<br>• Reflections and discussion of key points identified following workshop 2 |
| | KNOWLEDGE |
| | • Complete the ‘solutions in practice’ activity to establish how the SDM process could be introduced (by whom, when, where) and the supporting information required to enable stroke survivors to use the intervention independently, and supported by professionals (initially), caregivers and family/friends. |
| | EVALUATE |
| | • Summary of workshop lead by facilitator with group members invited to contribute (incl. feedback and questions), outline next steps and date of next meeting.<br>• Agree timescale and responsibility of members and researchers for contribute to the development of prototype intervention materials. |
| 4. How should we implement this intervention in clinical practice? What is ‘quality improvement’ and how do we use the QI principles to embed the process? If there | INFORM |
| | • Review evidence related to effective implementation of SDM<br>• Reflections and discussion of key points identified following workshop 3.<br>• Agree responsibility of members and researchers for specifying how the intervention should be introduced and implemented and the supporting information required to enable staff to introduce the SDM intervention and engage with patient and family in discussion about treatment options. |
| | KNOWLEDGE |

<table>
<tr><td rowspan="3">is documentation, where should this be stored? Do we need implementation groups within ward settings to embed this new intervention?</td><td><ul><li>Review prototype intervention materials developed following workshop 3.</li></ul></td></tr>
<tr><td>EVALUATE</td></tr>
<tr><td><ul><li>Participants to provide feedback on prototype materials.</li><li>Final revision of the prototype intervention, behaviour change strategies and implementation plan.</li><li>Recognition and celebration activity.</li><li>Summary of workshop lead by facilitator with group members invited to contribute (incl. feedback and questions), outline next steps.</li></ul></td></tr>
</table>

Table 2. quotes from interviews with family members and patients

| Theme | Quote number | Quotes |
|---|---|---|
| Experience of stroke and stroke care | 1 | P3: ‘’re; nursing care- couldn’t have asked for anything better’’ ‘’treatment was excellent’’ |
| | 2 | P2: ‘’don’t remember much of what happened in hospital’’. ‘’cant complain about the nurses’’ ‘’doctors were ok, but dealing with lots of patients and only spent certain amount of time’’ |
| Diagnosis and discussions about stroke and treatment, involvement in decision making | 3 | P1: ‘’3 different doctors, young doctor very good. Had a meeting in a room. Re: NG tube, ’I understand these things, he didn’t want it- he made it clear, he kept pulling it out’’. |
| | 4 | P2: ‘’shocked ...eh.... ’wasn’t in a position to make choices ‘’glad they (ref to doctors) made the choice- gave treatment’’ ‘’wasn’t aware of making any choices- people just came and went’’ ‘’you trust nurses and doctors to do what’s best for you- maybe its silly but I didn’t think’’ |
| | 5 | P4: ‘’ ’NG not advisable by doctor, asked if we wanted, neutral decision; we agreed not for as we know her- she wouldn’t have wanted that- would not want to prolong’’ |
| | 6 | P5: ‘’ ’it would be good to know when to speak to a doctor as doctors do rounds in the mornings’’ |

| | | |
|---|---|---|
| Provision of information | 7 | P3: ’information is a matter of choice whether people want it or not; maybe giving it later to ‘’taking it all in’’ |
| | 8 | P2: ’consultant and doctor came round with a machine and gave information’’ ‘’gave her some sort of idea of how serious’’ ‘’had leaflet, gave to son who came from France to look after her’’ ‘’info printed early on is fine, can look at it whenever’’ |
| | 9 | P3: ’good idea, give later on and would help, pictures would be good’ |
| | 10 | P5: ‘’ would have been useful. Other family can decipher. Useful information to know ongoing support and diagnosis. People remember things if there is a visual element to it- memory is triggered by a visual thing’’. |
